# Discretizing clinical information can reduce antibiotic misuse: a game theoretic approach

**DOI:** 10.1101/2020.08.23.20180117

**Authors:** Maya Diamant, Shoham Baruch, Eias Kassem, Khitam Muhsen, Dov Samet, Moshe Leshno, Uri Obolski

## Abstract

The overuse of antibiotics is exacerbating the antibiotic resistance crisis. Since this problem is a classic common-goods dilemma, it naturally lends itself to a game-theoretic analysis. Hence, we designed a model wherein physicians weigh whether antibiotics should be prescribed, given that antibiotic usage depletes its future effectiveness. The physicians’ decisions rely on the probability of a bacterial infection before definitive laboratory results are available. We show that the physicians’ equilibrium decision-rule of antibiotic prescription is not socially optimal. However, we prove that discretizing the information provided to physicians can mitigate the gap between their equilibrium decisions and the social optimum of antibiotic prescription. Despite this problem’s complexity, the effectiveness of the discretization solely depends on the distribution of available information. This is demonstrated on theoretic distributions and a clinical dataset. Our results provide a game-theory based guide for optimal output of current and future decision support systems of antibiotic prescription.

## Introduction

Resistance to antibiotics has significant public health implications worldwide^1,2^. Antibiotic resistant infections increase mortality rates, lengthen hospital stays and increase treatment costs^3^. When a patient is suspected of having a bacterial infection, bio-specimens are often collected, cultured and undergo antimicrobial susceptibility testing. Typically, the results are known within several days. Having the results, a physician can prescribe an appropriate “definitive” treatment. However, these days might be critical, and therefore an initial “empirical” antibiotic treatment is given before identification of the causative pathogen^4^. The choice of empirical treatment is based on the physician’s assessment of the probability of different types of infection, given the medical symptoms, clinical settings, and the results of immediate diagnostic tests.

Appropriate empirical treatment, that ex-post matches the culture results, is highly important. Mainly, it reduces the mortality rate due to bacterial infections significantly^5–7^. The obvious adverse effect of unnecessary empiric treatment is an excessive use of antibiotics, which drives the emergence of antibiotic resistant pathogens in the general population^1,8^, as well as in the treated patients themselves^9–11^, through the evolutionary forces of selection. Nonetheless, patients are often unnecessarily treated with antibiotics (i.e. when no bacterial infection is present)^12^.

The problem of resistance emergence has set a challenge to the medical decision-making literature, which traditionally focuses on the well-being of the individual patient. Both empirical and theoretical studies have been conducted to find antibiotic prescription strategies that decelerate the process of resistance emergence and spread^13–19^ and analyze its economic cost to society^20,21^.

Nearly none of the existing models have analyzed the *clinical* decision problem of initiating antibiotic treatment from a game-theoretic perspective but instead tried to identify the socially optimal policy – one that considers the benefit of both current and future patients. The underlying assumption in such models is that since antibiotics can only be consumed given the physician’s prescription, the recommended policy can *and will* be implemented, despite the inherent conflict between the social interest and the personal interest of a single patient. However, the assumption of compliance to antibiotic usage guidelines may be unwarranted. Physicians have been documented to ignore recommendations of support systems, both with regards to antibiotic therapy^22,23^ and in other settings^24–26^. These deviations may be analyzed as rational strategic behavior, using the framework of game theory.

A game-theoretic approach views a physician as a decision maker (player) who seeks the treatment policy that will optimize the expected utility of *his own* patients, given the behavior of the other players. The player is not assumed to consider the utility of patients who are not under his direct responsibility. This assumption can be interpreted in two possible ways: the physician may either act myopically, as if each patient himself were the decision maker, or he may act on behalf of both his *present* and *future* patients.

The first interpretation was recently modeled in^27^, but the model precluded physicians from considering any long-term implication of antibiotic usage on future patients. Furthermore, this model did not consider a potentially crucial factor underlying the clinical decision of whether to administer an antibiotic treatment: the differential information regarding the probability of bacterial infection in each patient. Hence, the modeled treatment strategies were limited to either treat/not treat everyone, or making a random decision.

The second interpretation of the game-theoretic approach has not yet been considered, and it is the main focus of our study. In this setting, the decision maker does not completely ignore the effect of antibiotic use on future patients, as in the case of myopic behavior. However, the decision maker does not completely comply to it as well, as in the case of socially optimal policy. Due to the abovementioned lack of compliance to decision support systems, the Nash equilibrium of this game may be viewed as a realistic representation of physicians’ decision-making. Here, the Nash equilibrium is a stable state where none of the physicians benefits from unilaterally changing her behavior. Therefore, it is important to find out under which conditions the equilibrium strategies correspond with the socially optimal policy. A significant difference between the physicians’ equilibrium and the optimal policy would lead to the conclusion that a full implementation of the social optimum should not rely solely on the “good will” of physicians.

This study analyzes the clinical decision problem of initiating an empirical antibiotic treatment from a game-theoretic perspective, and compares the socially optimal policy with the subgame perfect Nash equilibrium and the myopic policy. In the Methods section, the medical problem is described and modeled as a repeated game with imperfect information between two physicians in subsection 1, which also includes the description of the myopic policy. The social optimum and its main characteristics are analyzed in subsection 2 and the subgame perfect equilibrium in subsection 3, where we prove the existence of a symmetric subgame perfect equilibrium in which players use pure Markovian strategies, and show that the optimal policy and the equilibrium are never the same. Furthermore, we show that the equilibrium strategies always dictate an overuse of antibiotics, compared to the social optimum.

In the Results section, we propose a simple yet surprising solution to show that changing the structure of the data available to the physicians, specifically discretizing or “coarsening” it, can reduce the gap between a rational physician’s equilibrium and a socially optimal policy. The formal derivations and proofs of this result are given in subsection 4 of the Methods. This result is independent from any parameter values, apart from the shape of the physician’s information distribution. We show our results under different theoretic distributions as well as an empiric distribution we estimate from data of respiratory infections in children.

## Results

### The original model: continuous information signal

We describe a repeated (stochastic) game with imperfect information, in which physicians are “players”, who receive patients over time. Each physician decides whether to treat his current patient with antibiotics. The physicians’ decisions rely on a partial information signal about each patient: the probability of a bacterial infection before culture results are available. The single-patient decision problem of a physician is described in figure 1.

**Figure 1:**
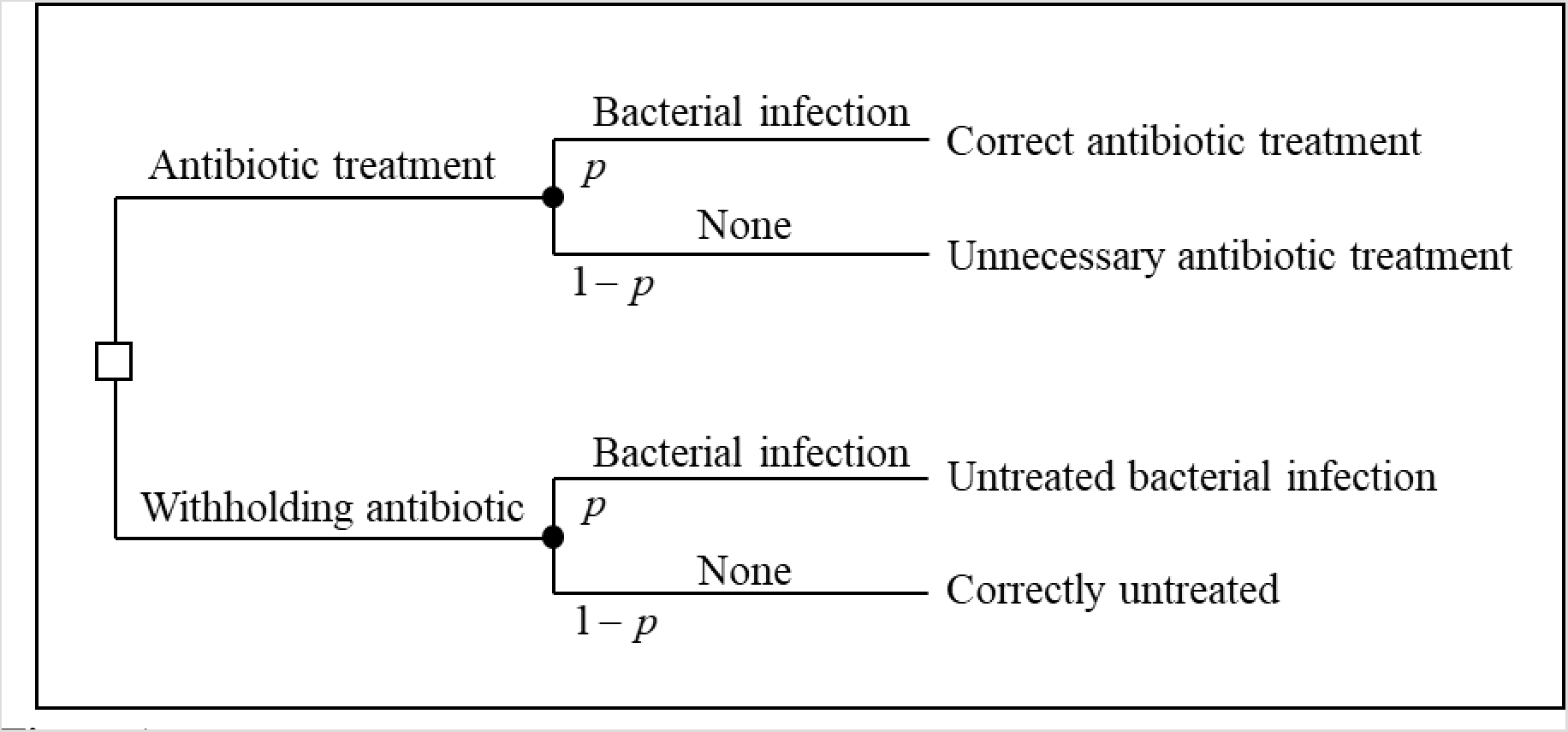
The tree of the single-patient decision problem of a physician given information signal *p*.

In addition, each use of an antibiotic drug depletes its future effectiveness due to antibiotic resistance. Thus, a strategy of the physician in our game describes the minimal probability of infection (threshold signal) that would make him administer antibiotic treatment at any given level of antibiotic effectiveness (“E-state” – the current sensitivity of bacterial population to the antibiotic). An extended description of the model appears in section 1 of the methods (1).

We then focused on contrasting two main treatment policies: A *socially optimal policy*, aimed to maximize the sum of *all* patients’ utilities over time; and an *equilibrium policy*, derived from each physician’s attempt to maximize the cumulative utility of all of *his own* patients over time, given the behavior of the other physician.

Under the social optimum, we show that as long as a “sufficient” amount of antibiotic effectiveness still exists (the E-state is large enough) - it is worth treating only patients with very high probability of infection (see theorem M.2.2 in section 2). When analyzing the equilibrium policy, we prove that a symmetric pure-strategy Markov Perfect Equilibrium (MPE) always exists (theorem M.3.4 in section 3.2). Moreover, under the *MPE the physicians always use antibiotics more extensively than the social optimum would dictate* (theorem M.3.5 in section 3.3)

### Implementing social optimality

We will try to eliminate, or at least reduce, the gap between the optimal policy and the MPE, by changing the “rules of the game”. The implementation problem^28^ is “the problem of designing a mechanism (game form) such that the equilibrium outcomes satisfy a criterion of social optimality”. The mechanism designer cannot impose the socially optimal policy on the players. Instead, he strives to set the rules of the game in such a way that the desired outcome will become an equilibrium. In our case, the possible changes in the rules of the game are constrained, mainly by ethics and norms. For instance, “fining” a player for using undesirable strategies is a common and effective means in mechanism design. However, as the payoffs in our game are based on survival rates or other measures of health, it is not possible to punish a physician by reducing his payoffs (i.e. harming his patients). In addition, it is not reasonable to make a clinical decision on the basis of the toss of a coin. Therefore, we will consider solutions that are based on *changing the information structure of the game*.

### Coarsening the information

Our goal now is to manipulate the information structure in a way that would turn the optimal policy into an MPE. Alternatively, in case this is impossible, we would like our manipulation to add a new equilibrium to the game, which is more socially desirable than the existing MPE.

The information system in our original model consists of each physician’s private signal, which we define as a continuous density function *f*(*p*) and which induces a posterior *p* that the patient has a bacterial infection That is, there is a probability of 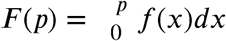 that a random patient has a posterior that is lower than or equal to *p*. This private signal can be, for example, the body temperature of the patient. The continuous signaling system can be replaced with a discrete signal system, which is less informative. The posteriors induced by the new signal are calculated using Bayes’ rule. We call this process “*coarsening* the information”.

If we have a continuous information system *f*(*p*) and we want to replace it with a dichotomous discrete system, we need to determine a certain threshold probability *T*. The new system contains two signals, high (*H*) and low (*L*), each of which appears in a certain probability and induces a different posterior that the patient has the infection (for detailed explanation and calculations of the relevant probabilities see section 4.1).

Notice that in the case of medical information, it is very simple to implement, especially when decisions are informed by predictive risk scores or probability assessments attained from algorithms – by receiving the results clustered into ranges instead of an exact number. For example, if the signal is the patient’s temperature, we can relay information only regarding ‘high’ Vs. ‘low’ temperature values, based on our selected threshold.

Similarly, if we want to replace the continuous information system with a discrete system that contains *J* signals rather than two, we will use a similar coarsening process with *J* − 1 thresholds. However, for the purpose of our discussion, it is sufficient to consider dichotomous signals.

### Information and Stability

By theorem M.3.2 we know that if any deviation in any direction (and specifically towards overuse of antibiotics) is worthwhile, then the minimal deviation in this direction is worthwhile as well (the *minimal* one-stage-deviation principle). However, the opposite claim is not necessarily true. In many cases it is possible that a physician has an incentive to perform a small deviation towards overuse, while a large deviation in the same direction would result in a loss.

Using a dichotomous signal system with a threshold *T* is equivalent to limiting the set of available decision rules in each E-state to either “treating only patients with a high signal” or “treating everyone”. Consequently, we limited the set of possible deviations from a recommended policy. Specifically, if the recommended symmetric policy is “treating only patients with a high signal”, a player may either deviate to “treating everyone” or stick to the recommendation. Thus, it is possible that this recommended policy will not be an equilibrium under the original continuous information system, and yet it will be an equilibrium under the dichotomous system.

As a result, although the optimal symmetric policy is never an MPE in a continuous information system, *we may be able turn it into one by coarsening the information* using a “very high” threshold. For example, let us assume once again that the signals are the patient’s body temperature. Suppose that a body temperature of at least 39°*C* induces a “very high” posterior that the patient has a bacterial infection (*p* → 1). Thus, the *optimal* policy is treating only patients with at least 39°*C*, and we want this policy to be an equilibrium. Assume a patient arrives with a body temperature of 38.9°*C*. The posterior probability that he has a bacterial infection is only slightly lower than that of a patient whose body temperature is 39°*C*, and thus the expected utility from treating this patient with antibiotics is rather high. Therefore, if the physician knows the exact body temperature of this patient, he has a strong incentive to deviate from the optimal policy. However, suppose now that we coarsen the information, and the new signal merely informs the physician whether the patient’s body temperature is above or below 39°*C*. If the physician only knows that his patient’s body temperature is in the range of 36 − 38.9°*C*, the new posterior that this patient has a bacterial infection is much smaller, and therefore the expected utility from treating him is much lower. Consequently, the physician’s tendency to follow the optimal policy will be much stronger.

When we use the new information system, the fixed symmetric policy of “treating only patients with a high signal” can be considered a good approximation of the original optimal policy. Therefore, we would like to know whether it can be an MPE.

Our main theorem states that ***the symmetric strategy combination, in which both players treat only patients with a high signal is an MPE if and only if***

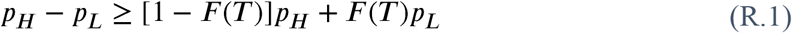

For detailed mathematical formulation and proof see theorem M.4.2 in section 4.3.

***Note that the condition in the theorem involves only the information of the physicians, and not any other parameter of the medical problem***.

The meaning of theorem M.4.2 is that treating only patients with a high signal is an MPE if the difference between the higher and the lower posterior of bacterial infection is not smaller than the prior probability of bacterial infection. This prior probability of a bacterial infection, *P*(*B*), is a given parameter of the original continuous information system *f*(*p*):

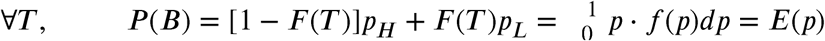

Which, conveniently, is the expected value of the posterior probability of a patient having a bacterial infection. Therefore, for any given continuous information system we can define *h*(*T*) = *P_H_* − *P_L_*, and find out what is the range of possible thresholds, subject to *h*(*T*) ≥ *E*(*p*).

From this theorem we can also conclude that if we specify the “symmetric strategy combination, in which both players treat only patients with a high signal MPE” as our goal, a coarsening process will fail only in either of the following “probabilistic” scenarios. First, when the prior probability of bacterial infection (the right side of equation (R.1) is very high to begin with. However, in this case, the damage caused by the instability of the optimal policy is limited, since even an overuse of the drug would yield a relatively high social utility. Second, if the original signal distribution has low dispersion resulting in a low difference between *p_H_* and *_L_* (the left side of equation (R.1).

## Signal distributions

To demonstrate the relations between the original information structure and the possible values of the threshold *T*, we present several possible scenarios of the signal distributions of a bacterial infection *f*(*p*) represented by a beta distribution with different parameter combinations (Figure 2.a-d). First, we present a scenario wherein often information regarding the infection is distinctly associated with a bacterial or non-bacterial disease (Figure 2.a). Such a scenario may arise when diagnosing endocarditis, where clinical information can be highly indicative of bacterial infection, rule it out almost certainly, or have some uncertainty^29^. In this case, we can set T at any value larger than 0 (Figure 2.e). In a scenario where patients are approximately as likely to arrive with different symptoms that define various degrees of certainty in the diagnosis of a bacterial infection, we can represent *f*(*p*) by a uniform distribution (Figure 2.b). Similarly, we can model information with uniform distribution on only a subset of the range of probability values, e.g. uniform on (0,0.5) and zero otherwise. A scenario pertaining to these kinds of distributions can be observed, for example, in pharyngitis diagnosis, for which a set of criteria have a range of probabilities attributed to them being indicative of a bacterial infection^30^. Then, once again, any threshold T can be turned into an MPE by coarsening the information (Figure 2.f). If most of the infections are non-bacterial, and *f*(*p*) is skewed to the right, we have some freedome in setting the value of T and can potentially reduce antibiotic misue by coarsening the signal given to physicians (Figure 2.c). This is similar to the empiric data of upper respiratory infections in young children, demonstrated in the next section. On the other hand, if in most cases a bacterial infection is approximately as likely as a non-bacterial one, i.e. *f*(*p*) is centered around 0.5, then there is not enough information in the system and no equilibrium T values can be set by coarsening the data (Figure 2.d).

**Figure 2:**
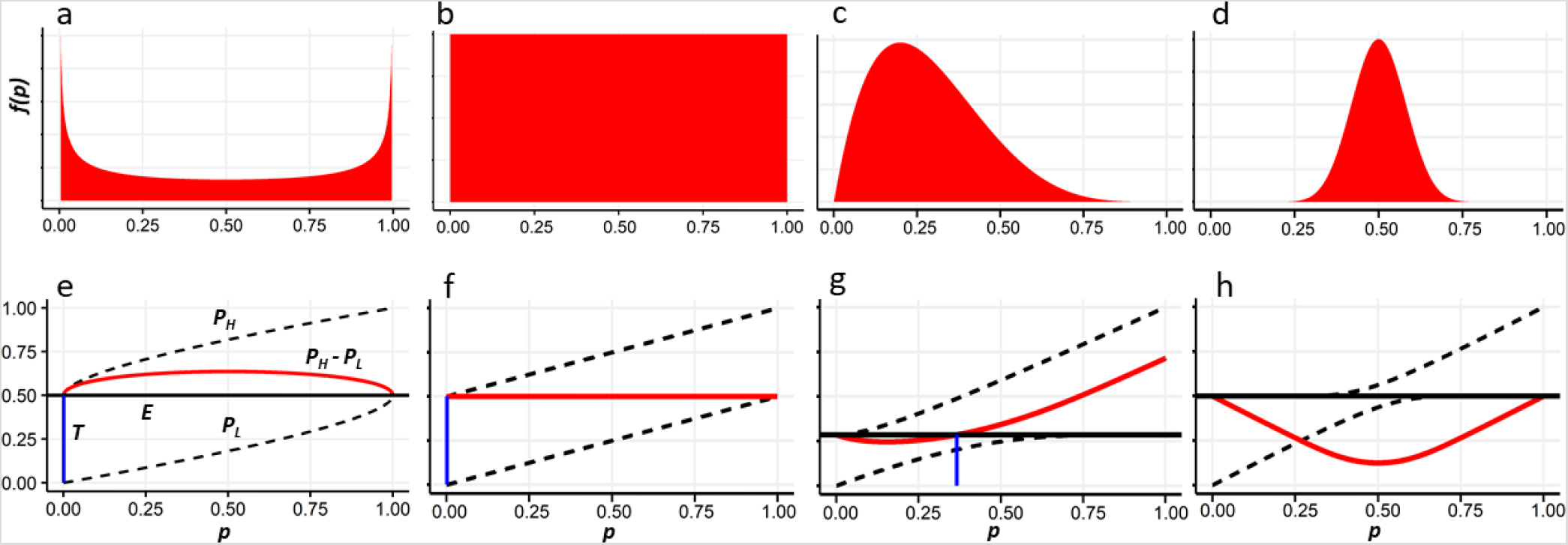
Theoretical posterior distributions of bacterial infection signals, and corresponding threshold values possible by coarsening the data. (a-d) Probability distribution functions *f*(*p*) representing distinct scenarios of a bacterial infection signal. (e-h) Corresponding (top-bottom) values of *E* (horizontal black lines), *P*_*H*_ and *P*_L_ (upper and lower dashed curves, respectively), their difference *P*_*H*_ − *P*_L_ (red curves) and the minimal threshold *T* (blue vertical lines) achievable by coarsening the data. All distributions are beta with parameters 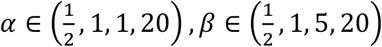, corresponding to panels a-d.

### Empirical example

We further investigated a real-world example. We obtained medical record data of 1202 children aged < 2 years who were hospitalized for a respiratory tract infection. We trained a machine learning model to classify bacterial and viral infections using the patients’ medical records, and used the model’s output as an estimate of the posterior probability of a bacterial infection – 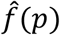 (Figure 3; see methods for details). For the obtained posterior distribution, coarsening the data can produce any threshold *T* ≥ 0.546, which can thus allow varying degrees of the strictness of antibiotic regulation upon a respiratory tract infection in children. That is, if the output of this toy model, or an improved machine learning algorithm, is provided to physicians to assist rapid classification of respiratory tract infections, a simplified output of ‘high’ and ‘low’ probability should help compliance and might mitigate antibiotic overuse.

**Figure 3:**
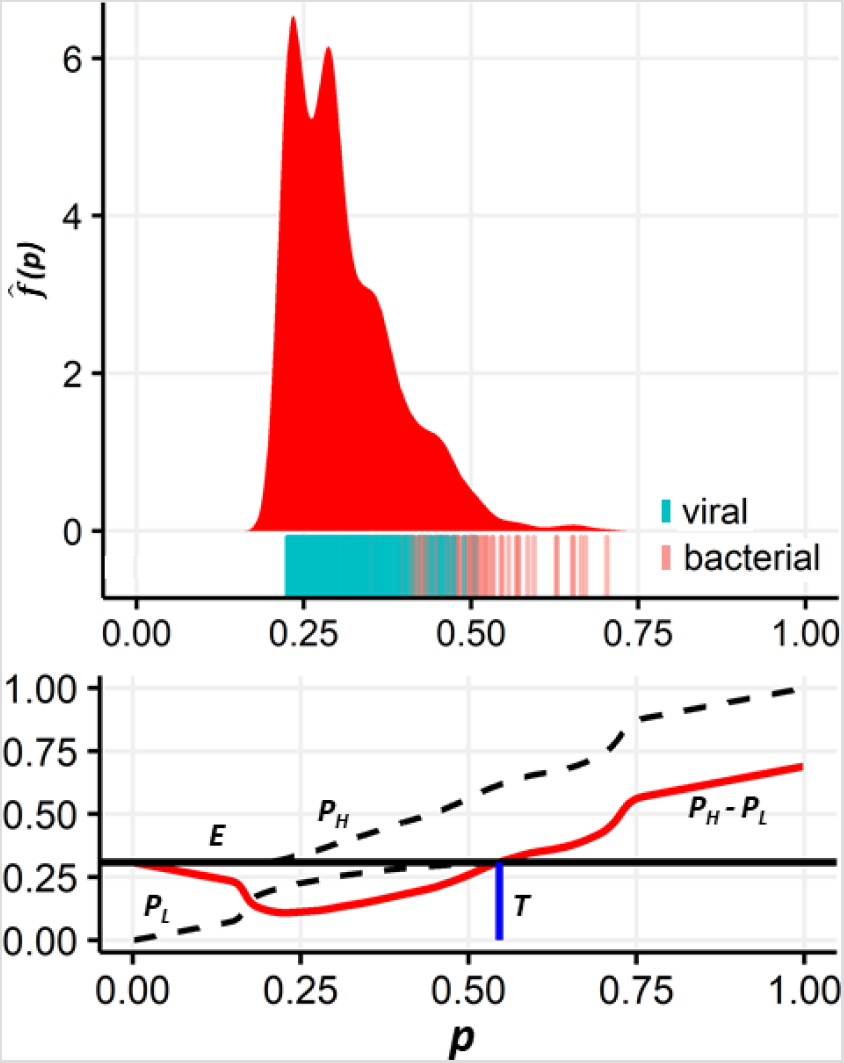
Posterior distributions of bacterial infection signals for children with respiratory tract infections, and the corresponding threshold values possible by coarsening the data. Top: The estimated probability distribution function *f*(*p*) associated with respiratory tract infections in children, caused either by a bacterial or viral (RSV) agent (see Methods). Under the distribution a rug-plot indicates the actual origin of infection in the data. Bottom: The corresponding values of *E* (horizontal black line), *P*_*H*_ and *P*_L_ (upper and lower dashed lines, respectively), their difference *P*_*H*_ − *P*_L_ (red curve) and the minimal stable threshold *T* (blue vertical line) achievable by coarsening the data.

## Discussion

This study applies the conceptual framework of game theory to analyze the conflict between individual and societal interests that is inherent to the decision problem of initiating an empirical antibiotic treatment. The problem is clear when the physician focuses on the well-being of a single present patient: in this case initiating antibiotic treatment is a dominant strategy. However, we have shown that the problem remains even if the physician does consider long-term implications, and cares about the well-being of his future patients.

We have shown that in this setting, although a symmetric SPE in pure Markov strategies always exists, it is not socially optimal, and the socially optimal policy is not an MPE. The analysis in section 3 of the Methods chapter, provides evidence that, although administering antibiotics to everyone is no longer a dominant strategy, the “rational” strategic behavior would lead to an overuse of antibiotics.

Our model reveals that in a setting that involves an interaction between several decision makers (physicians), supplying each of them with the maximal available information turns the socially optimal treatment policy into an instable recommendation (i.e., knowing the maximal information generates an incentive for each physician to deviate from the recommended policy). This is an unintuitive result, since in single-agent decision theory information cannot have a negative value, as it is either beneficial or ignored by the decision maker. We have further suggested a way to mitigate the problem by coarsening the information that is available to the physicians regarding the probability of a bacterial infection. Importantly, the conditions dictating when coarsening the information is socially beneficial depend solely on the distribution of the signal indicating whether the infection is of bacterial origin. This is a general result, independent of other parameters such as the utility of antibiotic treatment and antibiotic resistance frequencies.

Coarsening the information provided to a physician is already carried out in both this and other medical decision problems. For example, the Centor criteria help identify whether a sore throat is caused by a bacterial agent^30^, and the diagnosis is based on summing points allotted to symptoms and discretized age brackets. Similar discretizations of risk information are made for stroke^31^, endocarditis^29^, cardiovascular-related death^32^, pancreatic fistulas^33^, and many other medical applications. Our work shows for the first time that this discretization can guide “rational” physicians to more societally beneficial decisions.

In addition to the benefits of discretizing information presented in our results, providing detailed probabilistic information to physicians does not necessarily improve their decisions regarding antibiotic treatment. A study examining whether improving physicians’ judgments of the probability of streptococcal pharyngitis for patients with sore throats would affect their use of antibiotics, concluded that teaching physicians to make better judgments of disease probability may not alter their treatment decisions^34^. The authors hypothesized that even when the exact probabilistic information was available to physicians, they did not condition their decisions on it; but rather on ordinal judgments of disease probability or symptom severity, such as “not sick,” “mildly sick,” “moderately sick,” or “severely sick.” If this is the case, coarsened information can define these categories for physicians in a way that would benefit society. In other words, the goal of the coarsening is not to reduce the information available to the physicians but to simplify the decision-making process and thus make it more effective.

Although the analysis performed here is relatively general and relies on few assumptions, it naturally includes several limitations. Principally, the coarsening process may also add new undesired equilibria. In the worst case, it may even turn the myopic policy of treating everyone at every E-state into an equilibrium. However, a reasonable assumption is that a policy can be recommended to the physicians (either by some kind of authority or by a decision support system), and if the players have no incentive to deviate from this recommendation – it will indeed be accepted and implemented. Therefore, if the coarsening process indeed succeeds, the social interest in adding a “good” MPE outweighs the risk of adding a “bad” MPE.

We also assumed, for mathematical convenience, that the game ends when antibiotic effectiveness is completely depleted (E-state 0). In practice, administration of antibiotics with very high resistance frequencies is rare, but this has a negligible effect on our modeling framework. Low levels of drug effectiveness yield very low utility to the players, and therefore stopping the game at this point has no significant effect on their decisions until this point. Moreover, while we set the conditions for a successful information coarsening process (theorem R.2), we demanded that the new threshold T would be an equilibrium at any resistance level. If we, alternatively, specify a less “ambitious” MPE as our goal, e.g. “treating only patients with a high signal from E-state *k*′ onwards” (i.e., restricting the antibiotic treatment to patients with likely bacterial infections only when resistance levels are low), such a target MPE would only extend the range of possible T values.

Further limitations of our analysis is the focus on a decision including only the binary option of treatment/no treatment, and focusing on a two-player game between physicians. Regarding the former, a more general scenario should include different types of antibiotics, with different spectra of coverage and effect on resistance frequencies. In order to simplify the mathematical presentation, we have chosen to focus on the single antibiotic case, which contains the main aspects of the decision problem. Future research should also cover a wider framework, containing several drugs, e.g. representing prescription of broad or narrow spectrum antibiotics. However, this is a more complex case where the effect of one antibiotic on the other’s future effectiveness, due to cross-resistance^35–37^ between the different antibiotics, is not trivial and should be accounted for. With respect to the latter limitation, our results are relevant not only for single physicians. A “player” in a game theoretical model is often a group of people with common goals and utilities and centralized decision-making, such as a football team, a firm or even a country. In our case, a player may refer to a hospital or a country, in which many physicians apply the same clinical guidelines and policies. Consequentially our results can translate into the conflicts between the current and future patients of the specific hospital or country, rather than a certain physician.

To conclude, the problem of antibiotic misuse and resistance can be viewed as an example of “*the tragedy of the commons*”^38^. This concept has been previously used to describe the allocation of limited medical resources in scenarios of a conflict between individual and societal interests^39^. Previous studies called for collective solutions^40^, which may take the form of placing explicit resource constraints on resources available to physicians, or clinical practice guidelines that recognize cost-effective care as acceptable. The current study suggests another type of solution: supplying the physician with the level of information that would enable him to be committed to his patients without hurting society. This solution is expected to become increasingly relevant as data collection and computational power continually enhance the involvement of decisions support systems in medicine^41^ and specifically with respect to antibiotic resistance^42–44^.

Thus, by changing his available information, we allow the physician to act rationally and yet guide him towards the socially optimal decision and relieve him of the prisoner’s dilemma underlying the decision of antibiotic prescription.

## Methods

### 1 Formulation of the problem

The medical problem considered in this research involves a patient who has clinical symptoms that may indicate the existence of a bacterial infection. To simplify the analysis, we assume that only one type of antibiotic treatment is available, and that antibiotic treatment increases the chances of recovery of patients having bacterial infections while not affecting recovery of patients without bacterial infections.

The effectiveness of the antibiotic drug, denoted *e*, is the probability that the infecting bacteria are susceptible to it, whereas 1 − *e* will be the probability of resistance.

The effectiveness of the antibiotic drug is not a constant. Each use of the antibiotic drug (whether necessary or not) induces selection of resistant pathogens, and therefore reduces its future effectiveness (i.e. diminishes *e*). In addition, a patient who does not have the bacterial infection might acquire an opportunistic infection as a consequence of unnecessary antibiotic treatment. The chance of that happening is denoted *c*.

The physician, an agent acting on behalf of the patient, must decide whether an empirical antibiotic treatment should be administered. His decision relies on partial information about the patient: the probability of a bacterial infection in a given patient before culture results are available. This information is derived from symptoms, immediate diagnostic tests or decision support systems. The information signal results in a random variable parameter *p*, which is the posterior probability that the patient has the bacterial infection, where 0 ≤ *p* ≤ 1. The signal is a continuous random variable with a density function *f*(*p*) (see 1.2 under “information”). We assume the physician has complete information on all the parameters mentioned above, regarding the patient.

#### 1.1 Treatment Policy in the Static Model

Most of the medical decision-making focuses on the best interest of a single patient currently under treatment. Under this point of view, the question whether to administer an antibiotic treatment, given a certain signal, becomes a static decision problem. The static decision analysis takes the current level of effectiveness as a parameter, and ignores the current decision’s contribution to the emergence of resistance, and its effects on other future patients.

Following Pauker and Kassirer^45,46^, the static problem can be represented as a decision tree, and analyzed using the threshold approach derived directly from expected utility theory.

The aim of the static analysis is to calculate the “treatment threshold probability”, *T*. The optimal treatment policy of the physician is administering treatment if the probability of the patient having a bacterial infection exceeds the threshold. If we denote “Bacterial Infection” by *B* and “No Bacterial Infection” by *N*. The treatment threshold probability is calculated in the following way:

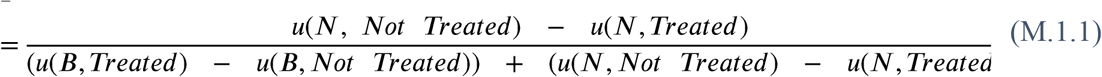

In the original analysis of Pauker and Kassirer, there are four possible outcomes, and each one of them is assigned a utility. However, as we have mentioned, in our case we assume that a patient who *does not have* the bacterial infection has the same chance of recovery with or without antibiotic treatment. Furthermore, we assume that since the case of bacterial infection involves a substantial risk, treating a bacterial infection is always preferred to not treating it, and not having a bacterial infection is preferred to having it, regardless of treatment. Thus, we can define three levels of utility: *r*_1_, the utility of “Bacterial Infection - Not Treated” (bacterial infection, without appropriate empirical antibiotic treatment); *r*_2_, the utility of “Bacterial Infection – Treated” (bacterial infection, with appropriate empirical antibiotic treatment); and *r*_3_, the utility of “No Bacterial Infection” (no bacterial infection, either with or without antibiotic treatment). Where *r*_3_ > *r*_2_ > *r*_1_ (note that the assumption regarding *r*_3_ is presented at this point only for the sake of compliance with the approach of Pauker and Kassirer, but it will not be needed later on in our complete model, from equation *(M.1.4)* onward).

For example, if we assume that the most important aspect of the possible outcomes is the survival of the patient, the utilities were assigned as follows: There are two possible states – “alive” and “dead”. Since we are dealing with Von Neumann-Morgenstern utilities^47^ we can arbitrarily set and *u*(*alive*) = 1 and *u*(*dead*) = 0. Consequently, the utility of each outcome is simply the conditional probability of survival. However, the utilities in the model can include other aspects, such as quality of life or financial costs, as long as the order of preferences is preserved (*r*_3_ > *r*_2_ > *r*_1_).

If we replace the final outcomes in the original tree with these utilities, denote *A* for “Administer Treatment” and *W* for “Withhold Treatment”, and add the posterior probability of disease induced by the patient’s symptoms (*p*), we get to the decision tree shown in Figure 4.a.

**Figure 4:**
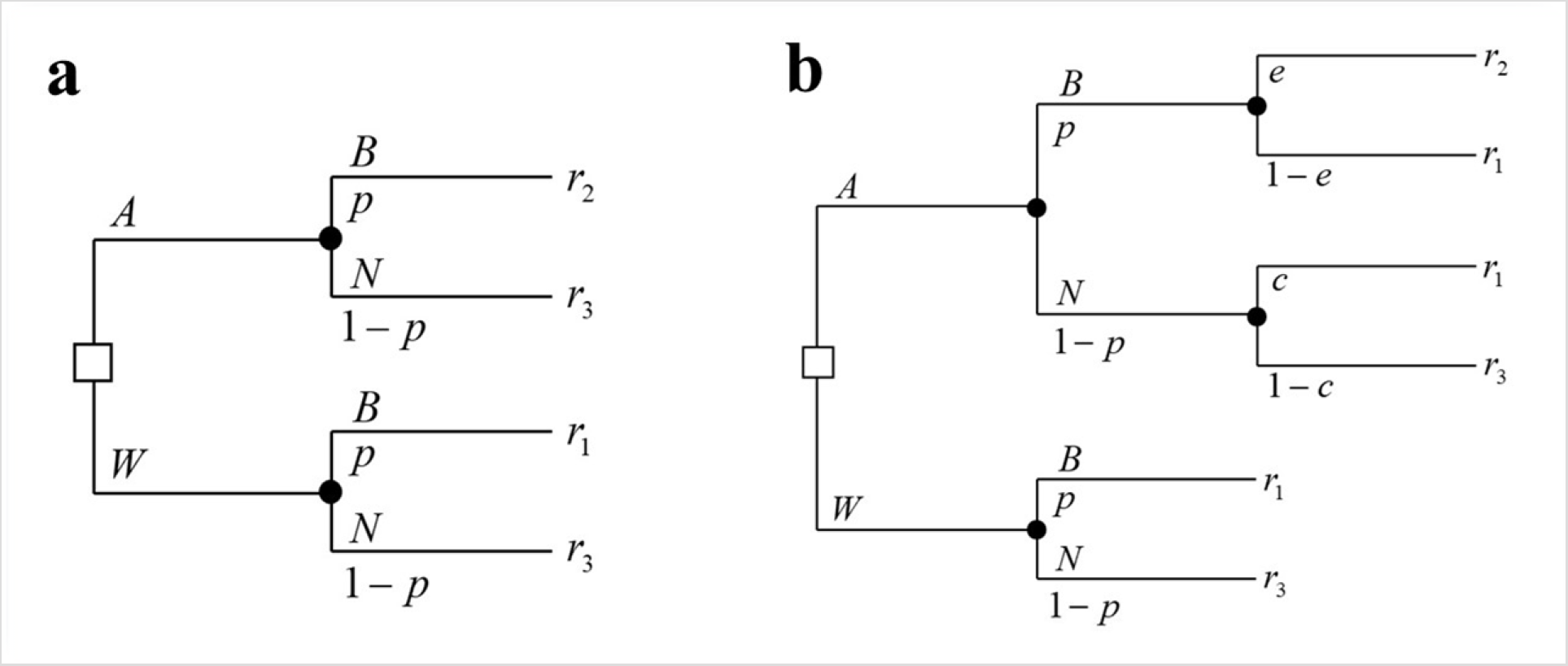
The decision tree of the single-patient static problem given signal *p*. (a) The basic decision tree including administering or witholding treatment (A and W, respectively), with the probabilities of bacterial and non-bacterial infections (B and N, respectively), and their associated utilities of the outcomes (r_1,_, r_2_, r_3_). (b) The tree extended to include antibiotic effectiveness (*e*) and the risk of unnecessary treatment (*c*). All variable definitions are given in Table 1.

However, two additional components of our model need to be included: the current level of drug effectiveness (*e*), and the risk that a patient who does not have the bacterial infection will develop one because of an antibiotic treatment (*c*). These two components have the form of additional “chance nodes” in our decision tree. Importantly, these do not add new type of outcomes, since they are simply “lotteries” between the worst outcome and a more preferable one. The decision tree of the single-patient one-period problem given a signal that induces a posterior *p* is shown in Figure 4.b. The definitions of all relevant variables are summarized in table 1.

**Table 1:**
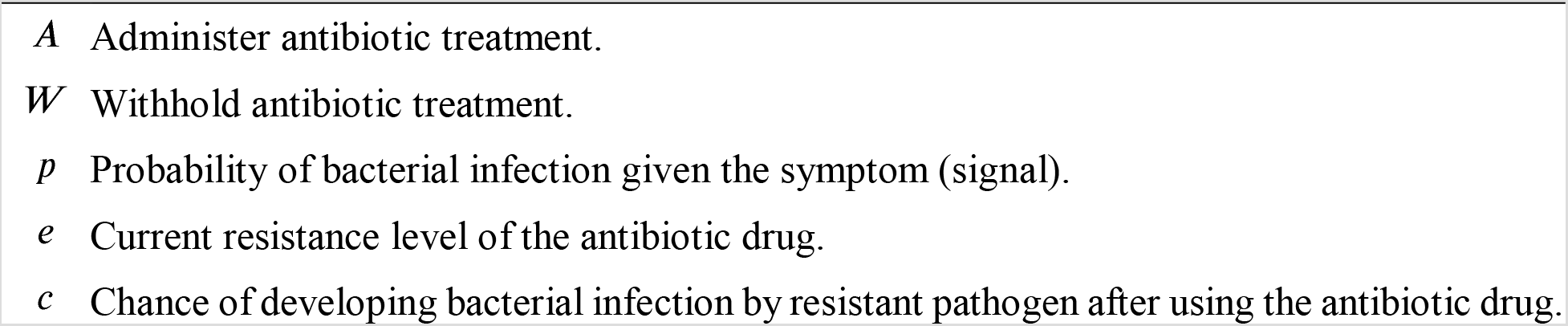
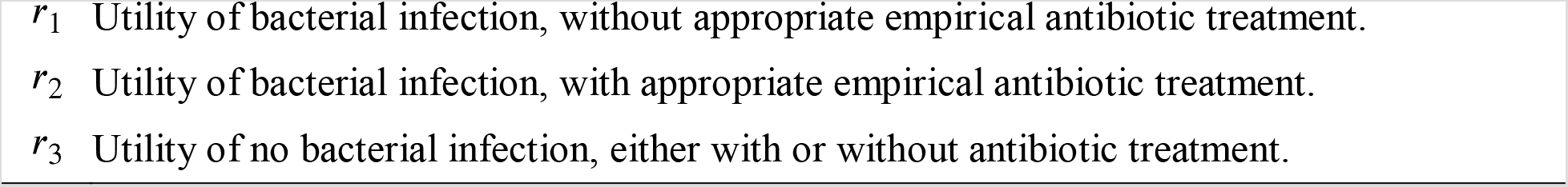
Definitions of the variables in the single-patient static problem.

|  |  |
| --- | --- |
| $A$ | Administer antibiotic treatment. |
| $W$ | Withhold antibiotic treatment. |
| $p$ | Probability of bacterial infection given the symptom (signal). |
| $e$ | Current resistance level of the antibiotic drug. |
| $c$ | Chance of developing bacterial infection by resistant pathogen after using the antibiotic drug. |

If we now want to calculate the threshold treatment probability in our model, equation *(M.1.1)* has to be adjusted in accordance with the new decision tree. The threshold *T* is defined as the probability (of bacterial infection) that generates indifference between administering and withholding antibiotic treatment. The optimal treatment policy of the physician is administering antibiotic treatment if the conditional probability induced by the patient’s signal exceeds the threshold: *T* ≤ *p*. The net risk of treatment is the loss of utility (or survival rates) caused by developing a bacterial infection as a consequence of the antibiotic treatment, multiplied by the chance of that happening. The net benefit of treatment is the net gain of utility (or survival rates) due to effective antibiotic treatment, multiplied by the chance that the treatment is indeed effective. The threshold treatment probability equation becomes:

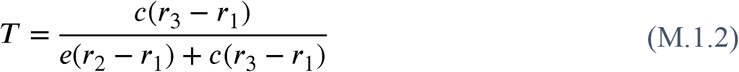

Note that unlike *(M.1.1)*, in the adjusted threshold equation (M.1.2) there are only three possible final outcomes (instead of four) and two extra multiplications: of the “net benefit” by the effectiveness coefficient *e* and of the “net risk” by the risk coefficient *c*.

#### 1.2 Treatment Policies in the Dynamic Model

Treatment policies in dynamic models involve the use of an antibiotic drug *over time*. In our dynamic setting, drug susceptibility (effectiveness) is regarded as a nonrenewable resource. This is a decent approximation of the state of antibiotic susceptibility – reversal of resistance is relatively slow and works on time scales bigger than resistance accumulation^48,49^. Therefore, any policy which aims to maximize the utility of different patients over time (either all patients or a selected group of them) has to take into consideration the future negative consequences of the antibiotic use. For the purpose of calculating social utility we will assume that it equals the sum of the individual utilities.

The dynamic model compares three types of treatment policies: *A myopic policy*, aimed to maximize the utility (or chance of survival) of the *current* patient at each point of time; *A socially optimal policy*, aimed to maximize the sum of all patients’ utilities over time; *An equilibrium policy*, derived from each physician’s attempt to maximize the cumulative utility of all of his own patients over time, given the behavior of the other physician.

Time in our dynamic model is discrete, each physician receives patients and makes decisions at discrete time points. The following definitions specify the characteristics of the game model.

##### Players

The basic setting is a game of two players (physicians) denoted 1 and 2, each of whom treats one patient in each period. A brief discussion of the extension to *n* players is given in section 1.3 and a more general interpretation of the two players model appears in the discussion.

##### Patient’s health condition

A patient either has a bacterial infection, *B*, or does not have it *N*. The patient’s condition is not known to his/her physician. We denote by *h_i_* the health condition of player *i*’s patient, *h_i_* ∈ {*B, N*}, *i* = 1,2. The prior probability of a patient having the bacterial infection in each period is *P*(*B*). These probabilities are independent within each period (between the two patients) and between periods (over time).

##### Actions

each player chooses one of two possible actions: to administer antibiotic treatment, *A*, or withhold antibiotic treatment, *W*.

##### Effectiveness depletion

As mentioned before, each use of the drug decreases the average antibiotic susceptibility to the drug, or its effectiveness, in the next period by a depletion effect *a*. The game ends when antibiotic effectiveness is completely depleted, for mathematical convenience. This assumption can be easily attenuated (see discussion). Given an initial effectiveness level *e*_0_ and the depletion effect *a*, one can calculate the *total number of possible game effectiveness-states*, *M*. Note that since the effect of use has a delay of one period, when the effectiveness is *α* two patients can be treated simultaneously before the effectiveness is completely depleted. Therefore, the number of patients that will be treated is either *M* or *M* + 1. Empirically, *M* is “very large” (an antibiotic drug is used by millions of patients before it is considered not effective), and for the purpose of mathematical convenience we will assume that it is as large as we want. For simplicity we will also assume that *e*_0_ = *Ma*, where *M* is a positive integer. The current level of effectiveness *e_t_* in any given period *t* is represented by the current *effectiveness-state* (E-state), 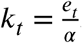. For simplicity, and since the E-state does not depend on the period number, we will omit the index and denote the E-state by *k*.

##### States and dynamics

The definition of state in our model has two aspects. As explained, the current E-state is defined by the current number of “doses” left, 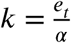. In each E-state the players face a “stage-game” with imperfect information. The information structure is defined in the next paragraph. In each stage-game there are four possible *health-states* (H-states), which are the combinations of the health condition of each of the current patients. The set of H-states is Ω = *H*_1_ × *H*_2_, where *H_i_* = {*B, N*} represents the health condition of player *i*’s patient. That is, for any fixed *k* the four possible H-states are: {(*B, B*), (*B, N*). (*N, BB*), (*N, N*)}. The system dynamically moves between states. The system dynamics has two components: deterministic and stochastic. The deterministic component is the transition between E-states, and it is determined by the player’s actions and the current E-state number *k*, through the effectiveness depletion. The stochastic component is related to the condition of the patients in the next period. It is stationary and depends on the distribution of patients, i.e. on *P*(*B*). Thus, with *k* > 1 doses left, the deterministic dynamics is moving from E-state *k* to *k, k* − 1 or *k* − 2 according to the actions described in Table 2.

**Table 2:** Transition matrix of the deterministic game.

|  |  | Player 2's action |  |
| --- | --- | --- | --- |
|  |  | A | W |
| Player 1's action | A | $k - 2$ | $k - 1$ |
| | W | $k - 1$ | $k$ |

The stochastic dynamics are:

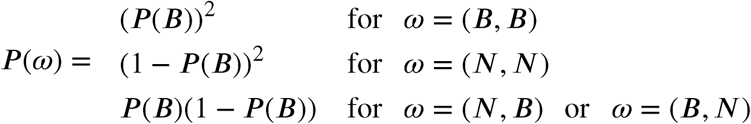

##### Information

We assume that each physician knows the E-state *k*. That is, physicians have information regarding the average resistance frequencies of bacterial infections in their cohorts. However, since the health condition of the patients is not known, the current H-state is not known by any physician. Each physician observes a signal regarding his patient. The information signal results in a random variable parameter *p*, which is the posterior probability that the patient has the bacterial infection, where 0 ≤ *p* ≤ 1. The parameter *p* is a continuous random variable with a density function *f*(*p*). We assume that *f* is integrable, and thus the probability that a patient’s posterior is *p*′ or less is 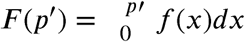. For simplicity, we will also assume that *f*(*p*) > 0, ∀*p*. This assumption can be easily omitted, by adjusting the strategy space, as explained later under “decision rules and strategies”. The patients’ signals within and between each period are independent. The signals are private information; each physician knows only his own patient’s signal. The distribution of the signals, *f*(*p*), is common knowledge. Note that the prior probability that a patient has the bacterial infection is: 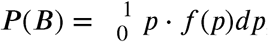, which is the expected value of the posterior distribution of a bacterial infection, later referred to as *E*(*P*).

##### The immediate payoffs

In each period these are defined as the *net* expected utility of antibiotic use, i.e. the difference between the utility of treatment and the utility of no treatment (otherwise, infinite utility can be accumulated by not treating any patient forever, due to the chance of a spontaneous recovery). Positive net utility (or gain in survival chances) can be achieved only if the patient had a bacterial infection and the treatment was effective. The probability of that happening, when signal *p_i_* is observed, is *p_i_* ± *e* = *p_i_* α *ka*. Negative utility (or loss of survival chances) is the consequence of developing infection as a result of the antibiotic treatment, when the patient did not initially have the infection. The probability of that event is *c* ± (1 − *p_i_*). Following are the immediate expected payoffs of physician *i* in E-state *k* when the signal of his patient is *p_i_*:

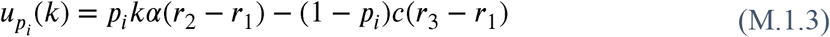

Notice that the immediate payoff of each physician depends only on his own action in the current period. The mutual effect is indirect and delayed, through the depletion of *e*.

##### Decision rules and strategies

A strategy of a physician is a mapping from histories (of E-states, H-states and actions) to actions. However, we are interested in a subgame perfect equilibrium in stationary Markovian strategies (also known as Markov perfect equilibrium - MPE). “Markovian” means dependence only on payoff-relevant variables, which in our model are the states (the E-states and the partial information about the H-states). “Stationary” means that these strategies will not depend on *t* (since time is not payoff-relevant in our model). In order to check that a combination of stationary Markovian strategies is an equilibrium, we need only to check that each player has no incentive to deviate to another *Markovian* strategy. The reason is that given any fixed stationary Markov strategy played by the other physician, the decision problem faced by physician *i* is equivalent to a Markov decision process (MDP)^50^. Thus, a best response exists in Markov strategies, and it can be found using a maximization process of dynamic programming. Therefore, we will denote strategies as Markovian, even though we do not actually limit a physician from deviating into a non-Markov strategy. A stationary Markov strategy is compounded of *decision rules*. A decision rule of a physician determines what to do in the *current* E-state, given the signal that *he currently* observes (the physician’s information about the current H-state), and not on the history. We will limit our discussion to threshold decision rules 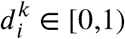, where physician *i* will choose *A* if 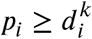 and will choose *W* if 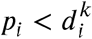. The decision rule 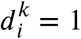 is not allowed, because it means not treating at all, since the probability that a patient has a posterior of *p_i_* = 1 or more is 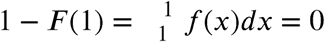. For the same reason, if we want to allow *f*(*p*) = 0 for *p_max_* < *p* ≤ 1 then we must limit 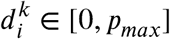. A pure (stationary Markov) strategy of physician *i* is, therefore, a vector containing the physician’s *M* decision rules for all the possible E-states 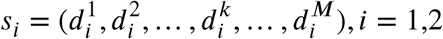. Note that the decision rules will be implemented in reverse order (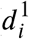 is the decision rule of player *i* with only one “dose” left to give, *e* = *a*). In addition, note that not all the rules will necessarily be applied in the realization of the game, because whenever both physicians choose *A* simultaneously the game skips E-state *k* − 1 and moves straight to *k* − 2. We denote by 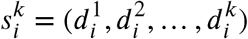 (the projection of *s_i_* on its first *k* coordinates) a strategy in the subgame that starts on E-state *k* (when 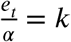) and continues onwards. We note that threshold decision rules are not only the intuitive, it is also easy to prove that they are the most efficient. The proof requires additional definitions that will be specified later on this section and appears in the supplementary information (S-3).

Finding the myopic policy is immediate. The emergence of resistance changes the net benefit of action (giving antibiotics) over time, but the future change has no influence on the decision in the current period. Therefore, the threshold probability is calculated in each period using *(M.1.2)* with the current level of effectiveness, *e_t_*.

However, in order to find the equilibrium policies and the socially optimal policy we need to analyze the repeated game characterized by the model. The socially optimal policy is derived assuming that one physician (a “social planner”) treats all the patients. Under this assumption, the game becomes a Markov decision process.

The information structure of the social planner problem matches the information structure of the game between the two physicians. Namely, the social planner is only allowed to condition his treatment policy on each patient’s signal, and not on the combination of signals. This setting enables us to compare the two cases, since it preserves the information limitations of the game, but it is also reasonable – in reality the number of both patients and physicians is much higher, and it is very unlikely to allow an action that depends on the simultaneous information of all. Following this line of reasoning we will also require that the socially optimal policy will be symmetric, i.e. the same decision rule (but importantly, not necessarily the same decision) is applied to both patients in every period.

The analysis of the game requires a more detailed definition of the payoffs and the concept of “deviation”. These definitions and the derivation of both optimal and equilibrium policies are described in the next section. For the sake of simplicity, and since is very small, we will assume from this point onwards that *c* = 0. Under this assumption, the net risk of treatment equals 0, and *using antibiotics is a dominant alternative* from the single-patient perspective. As a result, ***the myopic policy is necessarily to treat every patient***, regardless of his signal. The immediate expected payoff for a player who chooses action in E-state under this assumption is:

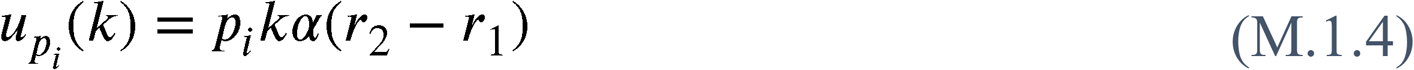

#### 1.3 Recursive Calculation of Payoffs

Payoffs can be calculated for any strategy profile 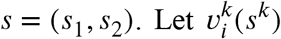. Let be the expected cumulative payoff of physician *i* from E-state *k* onwards, for a strategy profile *s*. The expected payoff is calculated with respect to the probability distribution of the signals. Note that the payoff depends on the strategies of the players only from this point onwards (i.e. their decisions for E-states 1,…*k*), and on the distribution of the future signals.

The value of 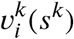 can be calculated recursively, starting from *k* = 1. The action taken by each physician in each E-state provides him with a certain immediate payoff, and the combination of actions taken by both physicians determines the future E-state, and thus their future payoff. Given the current E-state decision rule (threshold) of each physician, 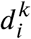, the probability that physician *i* will choose to administer treatment equals the probability that his patient’s signal will exceed the threshold, i.e. 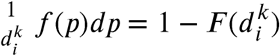. The different possibilities of expected payoffs on E-state *k* are represented in Figure 5. Note that the figure does not represent the game tree, but merely the four possible action combinations and their consequences.

**Figure 5:**
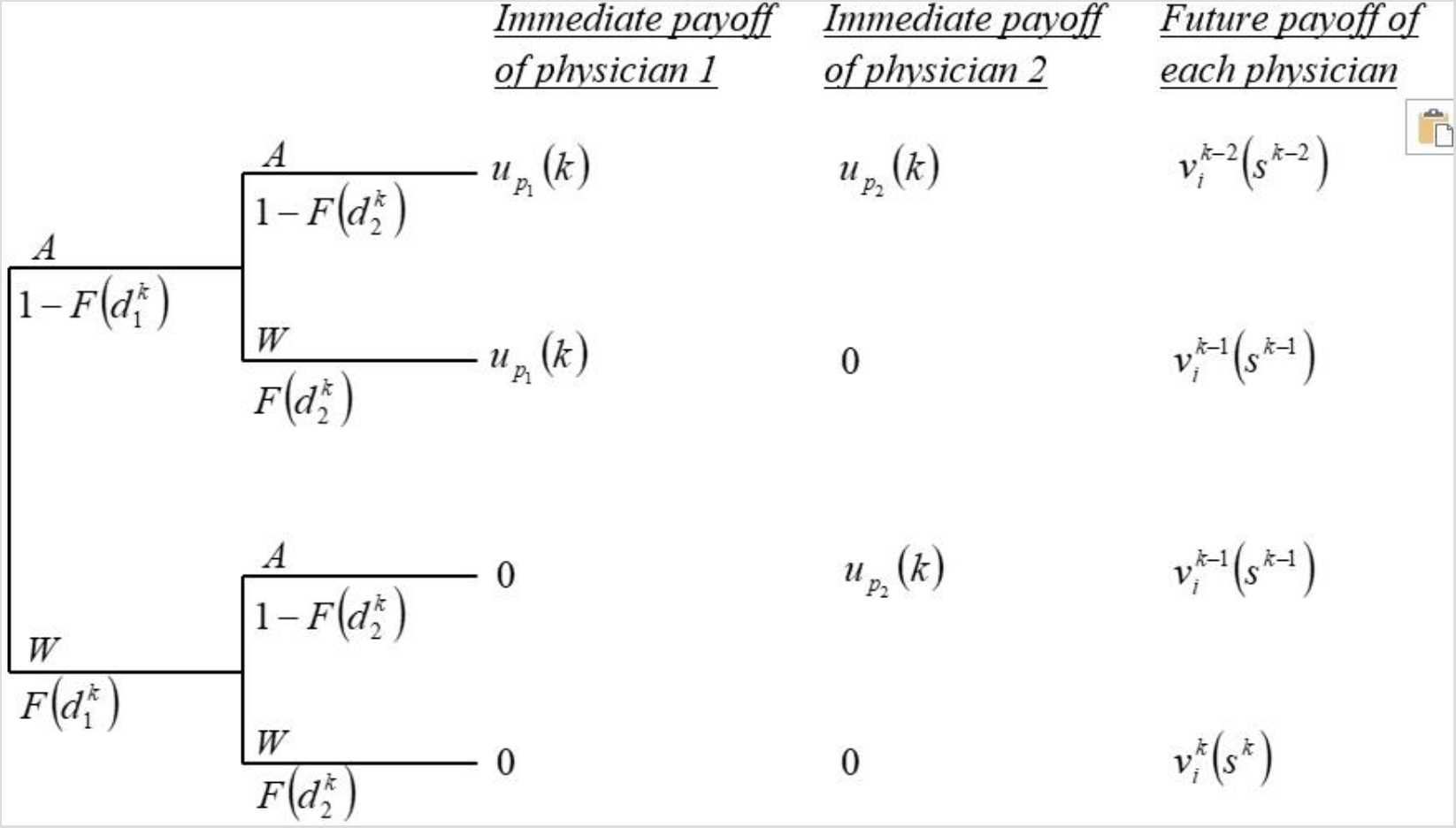
Expected payoffs of the players on E-state *k*.

**Table 3:**
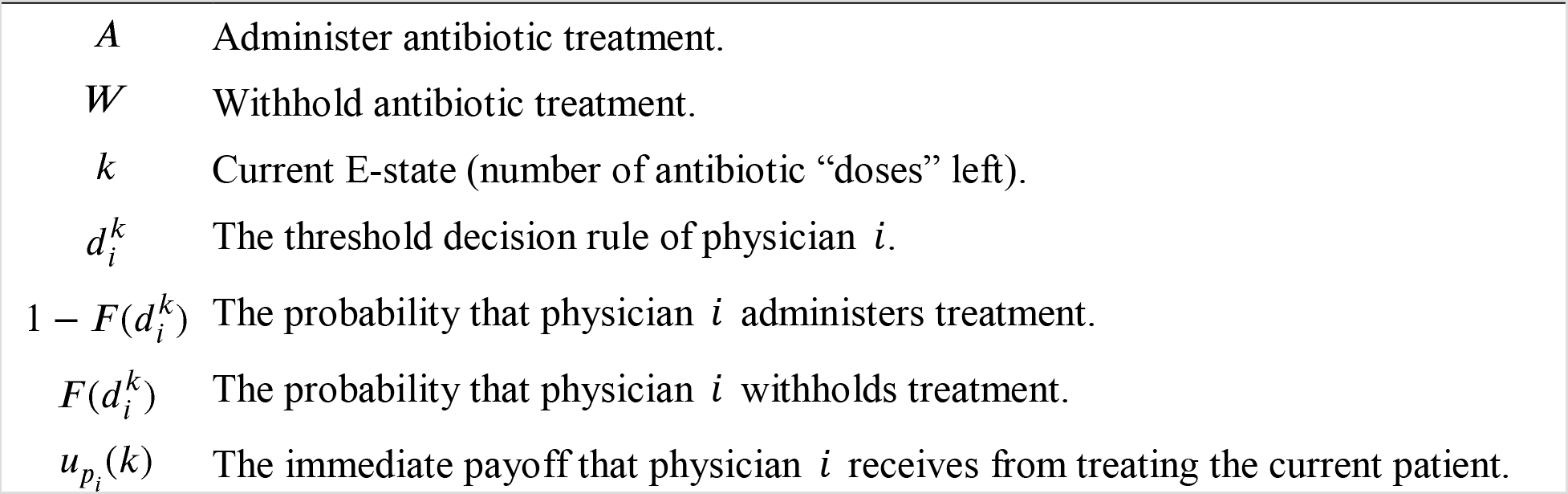
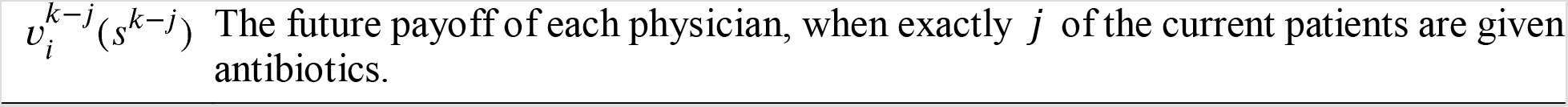
Definitions of the payoff variables in the dynamic game.

|  |  |
| --- | --- |
| $A$ | Administer antibiotic treatment. |
| $W$ | Withhold antibiotic treatment. |
| $k$ | Current E-state (number of antibiotic “doses” left). |
| $d_i^k$ | The threshold decision rule of physician $i$. |
| $1 - F(d_i^k)$ | The probability that physician $i$ administers treatment. |
| $F(d_i^k)$ | The probability that physician $i$ withholds treatment. |
| $u_{p_i}(k)$ | The immediate payoff that physician $i$ receives from treating the current patient. |

To facilitate the recursive calculation of payoffs we will sometimes write 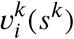 as 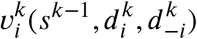. The payoffs can be calculated using the following recursive equation:

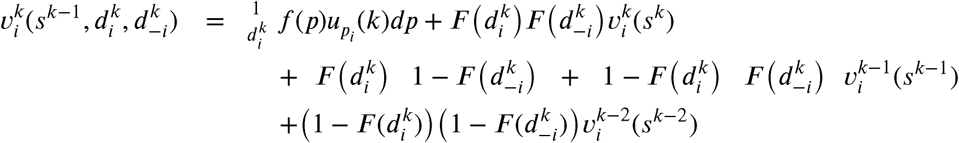

or, noting that 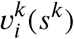 is on both sides of the equation,

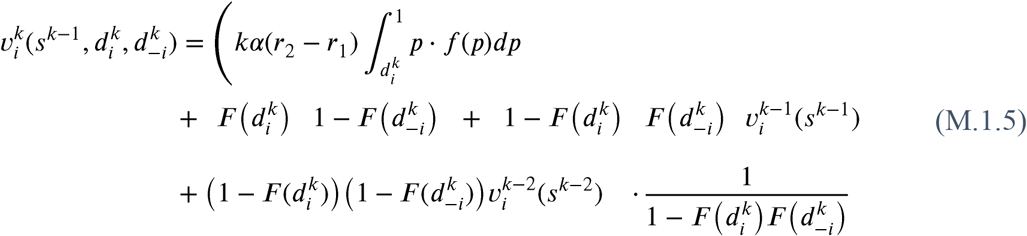

The numerator in *(M.1.5)* is the expected immediate payoff of player *i* given his own current E-state decision rule plus the expected future payoff given the strategy profile. The expected future payoff is the probability that either one or two physicians give antibiotic treatment, multiplied by the expected payoff with either one or two doses left, respectively. The denominator is the probability that at least one physician gives antibiotic treatment. For the sake of simplicity, we will sometimes use the following notation:

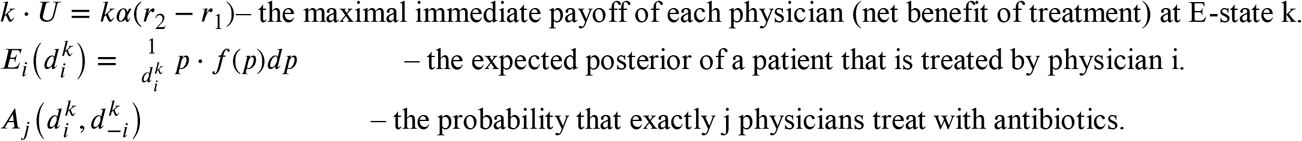

Specifically:

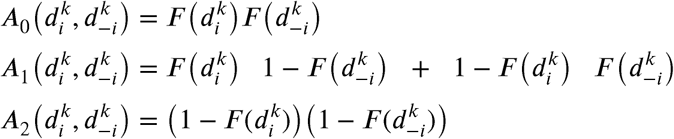

Using this notation, *(M.1.5)* can be written as

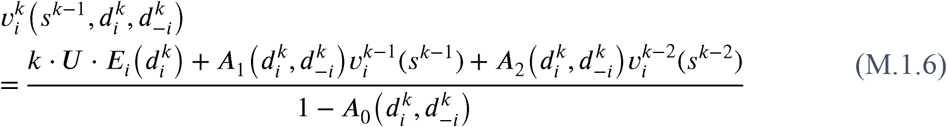

*Remark. (M.1.5)* can be easily extended to n physicians:

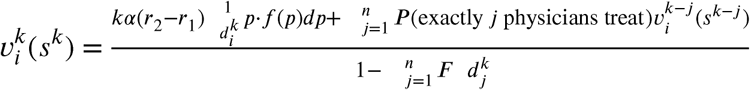

Note that since 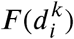 is continuous and 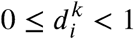, the payoff function 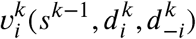 is also continuous. Due to the symmetry between the players and the symmetry of the social optimal policy, we will concentrate on searching for symmetric Nash Equilibria. Therefore, we will assume from this point onwards that the strategy profile is symmetric, i.e. 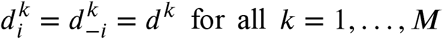 for all *k* = 1,… *M*. Under the symmetry assumption, *(M.1.5)* can be replaced by:

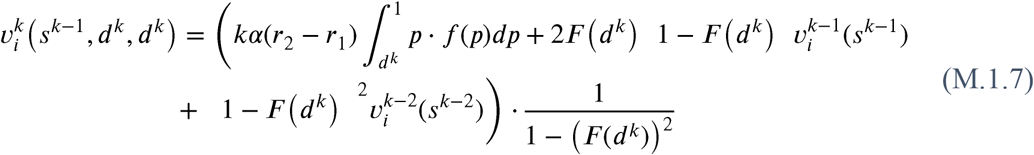

where 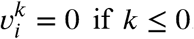.

The original equation of the non-symmetric case *(M.1.5)* will be used later on to check whether a player has an incentive to deviate at E-state from a given symmetric strategy combination.

### 2 The Social Optimum

In this section we will study the supremum of the payoff of symmetric policies. Though this supremum may not be obtained, we will describe how to approach it, and find an approximation of it.

Since the policies we are studying are symmetric, we will denote *s^k^* by the strategies of both physicians. Omitting the player index *i* we will look for the supremum of *v^k^*(*s^k^*) or, alternatively, *v^k^*(*s*^*k*−1^, *d^k^*). Let σ^*k*^ be the supremum of the payoff that can be achieved from E-state *k* onwards using a symmetric policy *s^k^*:

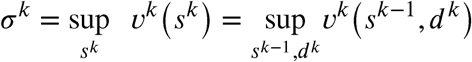

When we consider general policies, *d*^*k*^ = 1 is not allowed, because the payoff function can not be defined at 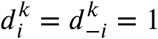, and the limit at this point does not exist. However, when we limit the discussion to symmetric policies, the definition of the payoff function can be extended to the case of *d^k^* = 1.

#### Lemma M.2.1

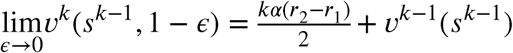

*Proof*. See supplementary information (S-4)

By lemma M.2.1 we can extend the symmetric payoff function for *d^k^* = 1, by defining

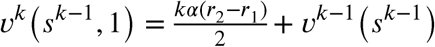

With this definition, the strategy space becomes a closed set, and the payoff function is continuous in it. Therefore, the extended payoff function has a maximum value. Let (*d*^1^, *d*^2^,…, *d*^k^,…) be a sequence of decision rules in the interval [0,1], such that *s^k^*, (*d*^1^, *d*^2^,…, *d*^k^) maximizes *v^k^*, that is, σ^*k*^ = *v^k^*(*s^k^*). Note that if this maximum value is achieved by strategies containing some *d^k^* = 1 then it cannot actually be obtained, but only approached.

The following theorem states the main characteristic of optimality in our model.

#### Theorem M.2.2

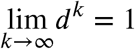

*Proof*. See supplementary information (S-5).

The intuition behind this theorem is that since the model is characterized by endless patience: as long as a “sufficient” amount of antibiotic effectiveness still exists - it is worth waiting for the patients with a very high probability of infection (“almost certainty”).

### 3 The Subgame Perfect Equilibrium

In this section we analyze the model as a non-cooperative game between the two physicians. Our goal is to compare the socially optimal policy to the equilibrium strategy of the game, and in particular to Markov perfect equilibria (MPE) of the game. In order for it to be comparable to the optimal policy we limit our discussion to pure symmetric equilibria of this type. In addition, based on our explanation of “decision rules and strategies” in section 1.2, when discussing the notion of “equilibrium” we need only consider individual deviations to Markov strategies, and not to history-dependent strategies.

This section consists of three parts: in section 3.1 we provide a criterion for a given policy to be an MPE, in section 3.2 we prove that such an equilibrium always exists and in section 3.3 we compare the MPE and the socially optimal policy.

#### 3.1 The Conditions for a Symmetric Pure-Strategy Markov Perfect Equilibrium

##### 3.1.1 The “One-Stage Deviation Principle”

The analysis of SPE in our model is based on the “one-stage deviation principle”. This principle is well known in the game theoretic literature; see for example Fudenberg and Tirole^51^: “in a finite multi-stage game with observed actions, strategy profile *s* is subgame perfect if and only if it satisfies the one-stage-deviation condition that no player *i* can gain by deviating from *s* in a single stage and conforming to *s* thereafter” (p. 109). An extended definition and explanation why this principle is valid in our model appears in the supplementary information (S-6).

Based on the “one-stage deviation principle”, at every E-state, *k* = 1,…, *M* the expected payoff under symmetric behavior (*d^k^*, *d^k^*) will be compared with the expected payoff of an individual stage-deviation from *d^k^* to *d^k^* ≠ *d^k^*. A symmetric strategy profile is an MPE if and only if

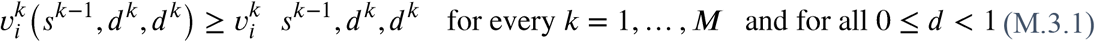

and we will use backward induction to verify this.

**Remark** *If the condition for profile is violated at E-state k* < *M, then any other strategy profile t with t^k^ = s^k^ is not an MPE as well*.

The condition *(M.3.1)* can be also represented as a set of difference-equations. The formulation of these equations will use the following definition:

For any given 0 ≤ *d^k^* < 1 let

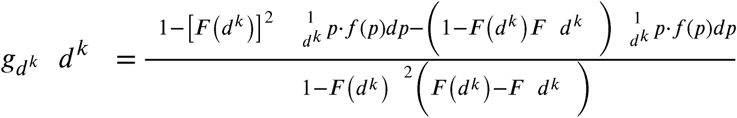

The formulation of the equilibrium equations using 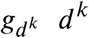 appears in the supplementary information (S-7).

The following Lemma states an important property of 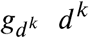.

###### Lemma M.3.1

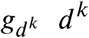 *is strictly increasing:*

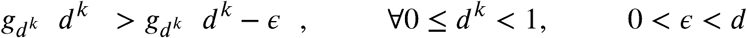

*Proof*. See supplementary information (S-8).

Following the last part of this proof, we define

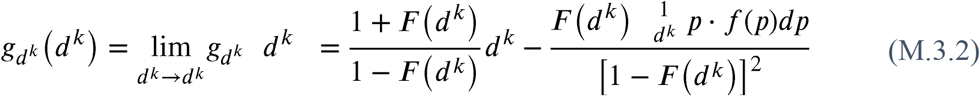

This definition will serve us later on in proving the gap between the socially optimal policy and the MPE.

##### 3.1.2 The Unimodality of Payoffs

The following theorem states that, when *k* > 1, for any given symmetric decision rule and assuming it is applied by the other physician, physician faces a unimodal E-state payoff function with respect to his own decision rule. Furthermore, the E-state payoff function is strongly unimodal, i.e. the global maximum is attained at a single point, and the function is strictly increasing until that point and strictly decreasing thereafter.

**Theorem M.3.2** *It is impossible that the following two conditions hold simultaneously at any given E-state k > 1:*

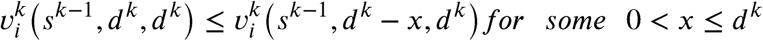

*and*

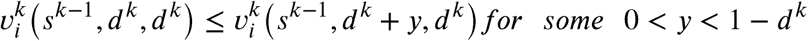

*Proof*. See supplementary information (S-9).

This theorem has two important implications for proving the existence of a pure symmetric MPE:

First, when we want to check if a physician has an incentive to deviate from a given symmetric decision rule in a certain direction, it is enough to check a very small deviation in that direction (there are no local maxima or inflection intervals). We shall term this notion the “Minimal One-Stage Deviation Principle”. Second, if a physician has an incentive to deviate from a given symmetric decision rule in a certain direction, then he necessarily has *no* incentive to deviate in the *other* direction.

#### 3.2 The Existence of a Markov Perfect Equilibrium in Pure Symmetric Strategies

Due to the “one-stage deviation principle”, in order to prove the existence of a symmetric pure MPE we can use backward induction. Starting from E-state *k* = 1, we only need to prove that at every E-state *k* there exists a symmetric pure “stage equilibrium”, 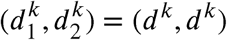. Theorem M.3.2 and the following lemma will enable us to do so.

##### Lemma M.3.3

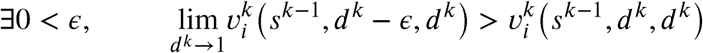

*Proof*. See supplementary information (S-10).

We can now prove the following existence theorem:

**Theorem M.3.4** *There always exists a symmetric pure-strategy MPE in the game*.

*Proof*. We can prove the existence of a symmetric pure-strategy MPE by constructing it, using backward induction. At each E-state *k* we will search for a symmetric pure-strategy “stage equilibrium” (*d^k^*, *d^k^*) given the symmetric pure-strategy equilibrium that was found in the previous E-states *s*^*k*−1^. Due to the “one-stage deviation principle”, if these symmetric pure-strategy “stage equilibria” exist for all *k* = 1,…,*M* (i.e. no player *i* can gain by deviating from *s* in a single stage) then there exists a symmetric pure-strategy MPE in the game.

The following process is performed at each E-state *k* = 1,…,*M*:

First, we check whether is a symmetric stage equilibrium. If it is - then an E-state-*k*-equilibrium exists (*d^k^*, *d^k^*) = (0,0), and we can move to E-state *k* + 1.

Otherwise - the physicians must have an incentive to deviate upwards (to *d_k_* > 0). Let

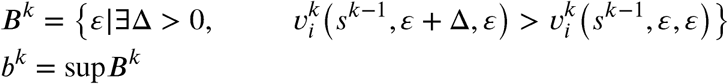

*B^k^* ≠ Ø because (0,0) ∈ *B^k^*.

*b^k^* ≠ 0 since the payoff function 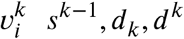 is continuous. Thus, if 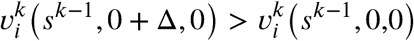 then there exist other symmetric combinations of decision rules close to from which each physician has an incentive to deviate upwards.

*b^k^* ≠ 1 because by lemma M.3.3 if is close enough to then each physician has an incentive to deviate downwards, and by theorem M.3.2 if a physician has an incentive to deviate downwards from a given symmetric combination (*d^k^*, *d^k^*) then he does *not* have an incentive to deviate upwards.

We now check whether (*b^k^*, *b^k^*) is a symmetric stage equilibrium. If a player has an incentive to deviate upwards, then due to the continuity of the payoff function there exists ∊ such that each player also has an incentive to deviate upwards from (*b^k^* + ∊, *b^k^* + ∊), but that contradicts the definition of *b^k^*. Similarly, if a player has an incentive to deviate downwards, then due to the continuity of the payoff function there exists Δ and an interval from (*b^k^* − Δ, *b^k^* − Δ) to (*b^k^*, *b^k^*) such that for every 0 > ∊ < ∊ a player has an incentive to deviate downwards from (*b^k^* − ∊, *b^k^* − ∊). But by theorem M.3.2, if at every (*b^k^* − ∊, *b^k^* − ∊) a player has an incentive to deviate downwards then he does not have an incentive to deviate upwards, and that also contradicts the definition of *b^k^*. *Therefore*, (*d^k^*, *d^k^*) = (*b^k^*, *b^k^*) *is a symmetric pure-strategy stage equilibrium*.

#### 3.3 Comparing the MPE and the Social Optimum

After proving that a symmetric pure-strategy MPE always exists, we would like to compare it to the social optimum that was analyzed in section 4. By theorem M.2.2 we know that as long as is large enough the social optimum is waiting for patients with a very high probability of a bacterial infection (“almost certainty”). We now show that the MPE is significantly different, namely that *in MPE the physicians always use antibiotics more extensively*.

**Theorem M.3.5** *Let* (*d^k^*, *d^k^*) *be a stage equilibrium*.

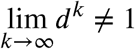

*Proof*. See supplementary information (S-11).

### 4 Reducing the Problem

After proving the existing gap between the optimal policy and the equilibrium of the game and the players’ rational incentive for overusing antibiotics, we strive to implement an approximation of the optimal policy as a new MPE of the game by coarsening the information available to the physicians. The motivation and practical interpretation of this process have been reviewed and explained in the results section. The following section contains its mathematical formulation and proofs of the main results.

#### 4.1 Coarsening the Information: Calculations

We now replace the continuous information system *f*(*p*) with a dichotomous discrete system signal system. We first need to determine a certain threshold probability *T*. The new system contains two signals, high (*H*) and low (*L*), indicating whether the patient’s probability of having bacterial infection is higher or lower than the given threshold *T*. Each of these two signals appears in a certain probability and induces a different posterior that the patient has the infection (see Table 4).

**Table 4:** Information signals and their probability.

| Signal | Probability of appearance | Posterior probability induced by the signal |
| --- | --- | --- |
| $H$ | $q_H = 1 - F(T) = \int_T^1 f(p)dp$ | $p_H = \frac{\int_T^1 p \cdot f(p)dp}{\int_T^1 f(p)dp} = \frac{\int_T^1 p \cdot f(p)dp}{1 - F(T)}$ |
| $L$ | $q_L = F(T) = \int_0^T f(p)dp$ | $p_L = \frac{\int_0^T p \cdot f(p)dp}{\int_0^T f(p)dp} = \frac{\int_0^T p \cdot f(p)dp}{F(T)}$ |

#### 4.2 Payoffs and MPE conditions with a Dichotomous Signal System

In order to explore the effects of the information coarsening on our game, we first need to adjust our definitions of payoffs (equations (M.1.5) and (M.1.7)) and MPE conditions (equation (S-7.1)) to a discrete dichotomous information system.

Briefly, the full model contains recursive calculations of payoffs. 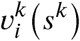 is the expected cumulative payoff of player *i* from E-state *k* onwards (from the current stage of antibiotic efficiency until the end of the game), for a strategy profile. It can be decomposed to 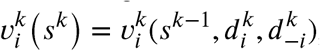, where 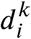 is the threshold of posterior probability of a bacterial infection defining the decision rule of player *i* in E-state *k*. The full description of the model and the definitions of all the variables appear in subsections Treatment Policies in the Dynamic Model and Recursive Calculation of Payoffs in the Methods section.

Using a dichotomous signal system is equivalent to limiting the set of available decision rules in each E-state to 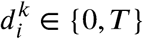. That is, there are only a low and a high signal.

Thus, there are only four possible payoff combinations in each E-state: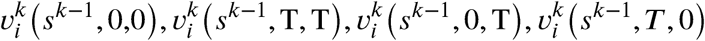.

For the extended calculations of the following equations see supplementary information (S-1).

If the strategy profile is symmetric, then the payoff of each player when they both treat everyone (*d^k^* = 0) is:

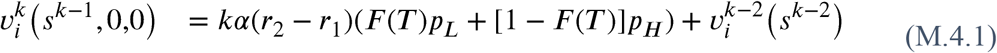

and when both treat only patients with a high signal (*d^k^* = *T*) it is:

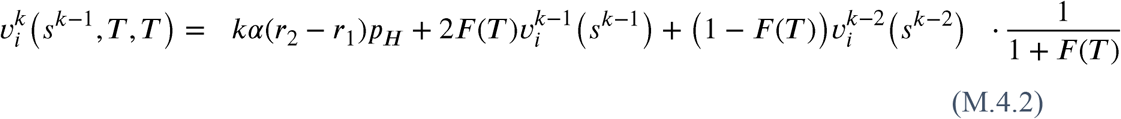

If the strategy profile is not symmetric, then the payoff of the physician who treats everyone is:

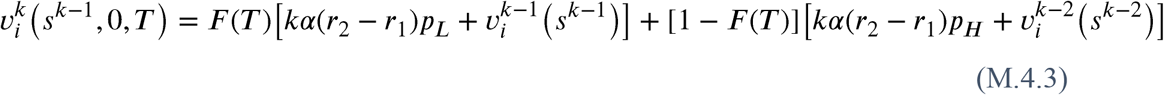

and the payoff of the physician who treats only patients with a high signal is:

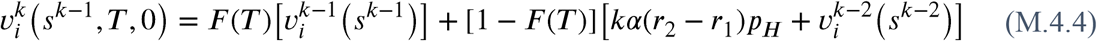

When we want to check whether a given *symmetric* strategy profile is an MPE, the “one-stage deviation principle”, wherein a strategy profile is an MPE if and only if no player can gain by deviating from it in a single stage and conforming to it thereafter^51^, still applies (as in our original information setting, see 3.1). However, there are only two types of possible deviations - either from *d^k^* = *T* to *d^k^* = 0 or vice versa.

Thus, *d^k^* = 0 (treating everyone) is a symmetric stage equilibrium if and only if there is no incentive to deviate to *d^k^* = T to (treating only patients with a high signal). Formally, iff:

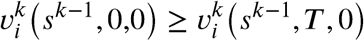

Using (M.4.1) and (M.4.4) and some algebra we get

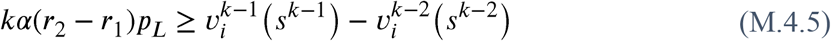

and, similarly, *d^k^* = *T* (treating only patients with a high signal) is a symmetric stage equilibrium iff:

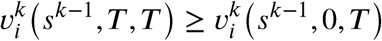

Using (M.4.2) and (M.4.3) and some algebra we get

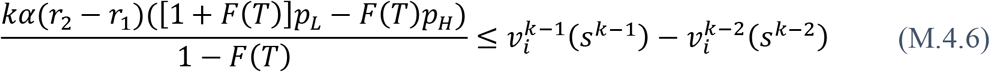

#### 4.3 Implementing Optimal Policy as a New MPE

When we use the new information system, the fixed symmetric policy of “treating only patients with a high signal” (i.e. *d^k^* = *T*, *k* = 1,…,*M*) can be considered a good approximation of the original optimal policy. Therefore, we would like to know whether it can be an MPE

In order to find the necessary and sufficient condition for this, we will use the following claim:

**Claim M.4.1** *Under the fixed symmetric policy s* = (*T, T*)^*M*^ (*i.e*. *d^k^* = *T*, *k* = 1,…,*M*)

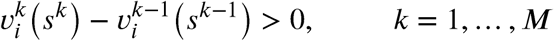

That is, the fixed symmetric policy of “treating only patients with a high probability of a bacterial infection” yields a total expected payoff (from E-state *k* onwards) that is strictly increasing in *k*.

*Proof*. See supplementary information (S-2)

Now we can use (M.4.6) and claim M.4.1 to set the following theorem.

**Theorem M.4.2** *Treating only patients with a high signal, s = ((T,T)^M^), is an MPE iff*

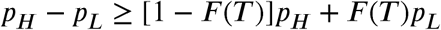

*Proof*. Due to the “one-stage-deviation principle” and (R.6) *s* = ((*T,T*)^*M*^ is an MPE if

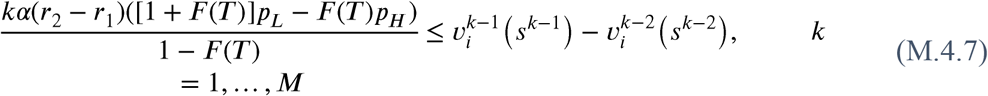

When *k* = 1:

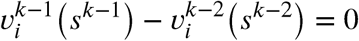

and by claim M.4.1, when *k* > 1:

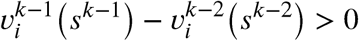

Therefore, if we want (R.7) to hold for all *k* = 1,…,*M* we get

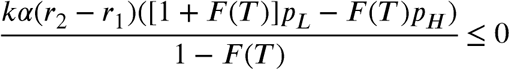

which means

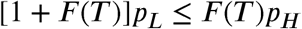

and finally

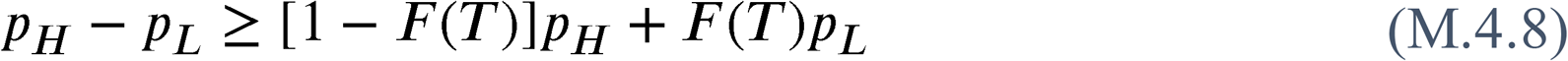

## 5 Data description and analysis

We obtained data of 1202 children aged < 2 years were hospitalized for bronchiolitis at Hillel Yaffe Medical Center in Hadera, Israel between 2008-2018. All children tested positive for RSV by antigen detection enzyme immunoassay, but 967 had only RSV bronchiolitis (viral infection), whereas 235 also had bacterial pneumonia, as confirmed in an in an X-ray scan. After retaining only variables with < 10% missing values, our data contained 27 patient variables, including demographics (e.g. age, sex, place of birth), clinical symptoms and signs (e.g. temperature, tachypnea etc.), comorbidities and the season of hospitalization. Values were imputed using a random forest algorithm^52^. We used the patient data to train a gradient boosted tree model^53^ that classified bacterial and viral infections. Briefly, we tuned the hyper parameters of the model using a 10-fold cross validation and applied the model to the entire dataset to recover for each patient a value 0 < *p* < 1. The estimated distribution of 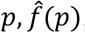, was a smoothed version of the resulting distribution of the patients, with a small positive constant (10^−4^) added to the distribution to account for sampling limitations and create a support of (0,1). This 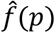 was considered as an approximation for the posterior distribution of having a bacterial infection. The final model output performed well with an area under the receiver operating curve (AUC) of 0.81.

## Data Availability

Data are proprietary.

